# Implementation of national guidelines on antenatal magnesium sulfate for neonatal neuroprotection in England, Scotland and Wales: Extended evaluation of the effectiveness and cost-effectiveness of the National PReCePT Programme

**DOI:** 10.1101/2024.07.16.24310419

**Authors:** Hannah B Edwards, Carlos Sillero-Rejon, Hugh McLeod, Elizabeth M Hill, Brent C Opmeer, Colin Peters, David Odd, Frank de Vocht, Karen Luyt

## Abstract

**Background:** Since 2015, UK national guidelines have recommended antenatal magnesium sulfate (MgSO_4_) for mothers in preterm labour (<30 weeks’ gestation) to reduce the risk of cerebral palsy in the preterm baby. However, implementation of this guideline in clinical practice was slow, and MgSO_4_ use varied between maternity units. In 2018, the PReCePT programme, an evidence-based Quality Improvement intervention to improve use of MgSO_4_, was rolled-out across England. Earlier evaluation found this programme to be effective and cost-effective over the first 12 months. We extended the original evaluation to determine the programme’s longer-term impact over four years, its impact in later preterm births, impact of the COVID-19 pandemic, and to compare MgSO_4_ use in England, Scotland, and Wales.

**Methods:** Quasi-experimental longitudinal study using data from the National Neonatal Research Database on babies born <30 weeks’ gestation and admitted to an NHS neonatal unit. Primary outcome was the proportion of eligible mothers receiving MgSO_4_, aggregated to the national level. Impact of PReCePT on MgSO_4_ use was estimated using multivariable linear regression. The net monetary benefit (NMB) of the programme was estimated.

**Results:** MgSO_4_ administration rose from 65.8% in 2017 to 85.5% in 2022 in England. PReCePT was associated with 5.8 percentage points improvement in uptake (95%CI 2.69 to 8.86, p<0.001). Improvement was greater when including older preterm births (<34 weeks’ gestation, 8.67 percentage points, 95%CI 6.38 to 10.96, p<0.001). Most gains occurred in the first two years following implementation. PReCePT had a NMB of £597,000 with 89% probability of being cost-effective. Following implementation, English uptake appeared to accelerate compared to Scotland and Wales. There was some decline in use coinciding with the onset of the pandemic.

**Conclusions:** The PReCePT Quality Improvement programme cost-effectively improved use of antenatal MgSO_4_, with benefits to the babies who have been protected from cerebral palsy.

**What is already known on this topic:**

- Antenatal magnesium sulphate (MgSO_4_) reduces the risk of cerebral palsy in babies born preterm.
- The National PReCePT Quality Improvement Programme (NPP) effectively and cost-effectively improved use of MgSO_4_ in England in the first 12 months of implementation, but sustaining quality improvements over time is often challenging.

**What this study adds:**

- Using a quasi-experimental design and routinely collected, longitudinal, patient-level data, this study found that the NPP had sustained effectiveness and cost-effectiveness over four years following implementation.
- Improvement may have been accelerated in England, compared to Scotland and Wales, where the NPP was not formally implemented.

**How this study might affect research, practice or policy:**

- This study demonstrates that dedicated national programmes can cost-effectively achieve improvements in perinatal care. The PReCePT model could be used as an implementation blueprint for other quality improvement initiatives in perinatal care.

## INTRODUCTION

Since 2015 the World Health Organisation (WHO)(1) and the UK National Institute for Health and Care Excellence (NICE)(2) have recommended administration of magnesium sulfate (MgSO_4_) in preterm deliveries <30 weeks’ gestation as a core part of maternity care. This follows strong evidence that when given antenatally to women in preterm labour, MgSO_4_ reduces the risk of cerebral palsy (CP) in preterm babies by around 30%(3). Historically, use of this treatment has been inconsistent, with only 64% of eligible women in England being treated in 2017. High regional variation in uptake also indicates inequalities in perinatal care(4).

As well as the significant impact of CP on affected individuals and their families(5), there are lifetime societal costs of approximately £1m per affected individual(6), and £1.8 billion annually on NHS clinical negligence litigation (half of the total NHS litigation expenditure)(7). Incidence of CP has been estimated at around 1.5 per 1000 livebirths in the UK(8), with preterm birth as the leading risk factor(9–11). This highlights the importance of funding effective, and cost-effective, strategies to reduce the risk of CP associated with preterm birth. It is estimated that one case of CP can be prevented, below 30 weeks’ gestation, for every 37 mothers treated with MgSO_4_, and around 200 cases of CP per year could be avoided by consistent administration of MgSO_4_ during labour(3).

In 2018, NHS England rolled-out the National PReCePT (Prevention of cerebral palsy in preterm labour) Programme (NPP). This was a quality improvement (QI) programme for maternity units, providing clinical guidance, training, learning resources, midwife backfill funding, and QI support, to improve maternity staff awareness, and increase use of MgSO_4_ for mothers in preterm labour. The aim was to reach ≥85% uptake in eligible mothers across all maternity units in England. The programme was delivered by regional Academic Health Science Networks (AHSNs, now Health Innovation Networks). Evaluation of the first 12 months of the programme found it to be effective, improving MgSO_4_ use by an estimated 6.3 percentage points, with an estimated net monetary benefit of £866 per preterm baby, and >95% probability of being cost-effective(12). However it is unknown whether these improvements were sustained over time, and sustainability in large scale implementation programmes is often a problem (and measurement of sustained effect often neglected)(13).

The primary aim of this study, therefore, was to evaluate the NPP’s longer-term, sustained effectiveness and cost-effectiveness over the first four years following implementation. A key secondary aim was to explore the impact of the NPP on all babies born up to 34 weeks’ gestation; NICE guidelines recommend treatment for births up to 30 weeks’, and ‘consideration of treatment’ for older preterm births up to 34 weeks’. Other secondary aims were to explore the impact of the COVID-19 pandemic on MgSO_4_ use, and to compare MgSO_4_ use in England with that in the devolved nations Scotland and Wales. This evaluation is part of a larger programme of work, including qualitative interviews to explore how the devolved nations were responding to the NICE guidance, reported elsewhere(14).

## METHODS

### Design

This was a quasi-experimental study for the evaluation of the NPP’s effectiveness and cost-effectiveness. The pre-registered Statistical Analysis Plan and Health Economic Analysis Plan were uploaded to the Open Science Framework prior to analyses: https://osf.io/be76s/.

### Intervention

The intervention being implemented was the National PReCePT Programme (NPP), as described above and fully detailed elsewhere(12), for the adoption of MgSO_4_ as a neuroprotectant in preterm births.

### Setting

NHS maternity units in England, Scotland and Wales. Within maternity units, analysis was performed on aggregated data on babies born preterm <30 weeks’ gestation, and admitted to an NHS neonatal unit, between January 2014 to December 2022. All maternity units in England, Scotland and Wales were included, excepting the five units in England that took part in the original PReCePT pilot study(15) and were therefore not part of the NPP.

### Data sources

Data on eligible babies and their mothers were obtained from the National Neonatal Research Database (NNRD), which holds individual-level, pseudonymised, routinely collected patient data on babies admitted to an NHS neonatal unit. Costs associated with the NPP were estimated in the original evaluation(12).

### Effectiveness evaluation

#### Outcome

The main implementation outcome was MgSO_4_ uptake over time. MgSO_4_ uptake was defined as the proportion of eligible mothers recorded as receiving MgSO_4_ at a maternity unit, expressed as a percentage. For consistency with nationally reported data, mothers with missing MgSO_4_ data were excluded from this calculation. This was computed per month per unit, and reported as a percentage. For consistency with nationally reported data only data on singletons and the first born (i.e. one infant) from each multiple birth were included in the calculation.

#### Descriptive analysis

Maternity unit, population characteristics, and MgSO_4_ use were descriptively reported by nation and time-period.

#### Primary analysis

Primary analysis was an interrupted time series using English data aggregated to the national level (mean national MgSO_4_ uptake per month, across all English maternity units). A multivariable linear regression model was used to estimate the difference in mean MgSO_4_ uptake from before (the 1-year period before) to after (the four-years follow-up) implementation of the NPP in England. The model adjusted for an underlying linear time trend, and mother and baby characteristics aggregated nationally per month (mean maternal age, Index of Multiple Deprivation (IMD) decile(16), baby’s birthweight adjusted for gestational age as a z-score, and proportion reported smokers, white British ethnicity, type of birth (c-section versus vaginal delivery), and multiple births. The model was further adjusted for a non-linear temporal component to account for the ceiling effect and reduction in rate of change at levels close to the ceiling. Data on paternal age and ethnicity were explored as potential confounding factors, but were excluded due to high levels of missing data, and expected collinearity with other variables (maternal and paternal age tends to correlate, as does maternal and paternal ethnicity). Potential interaction was explored between mean MgSO_4_ uptake and level of maternity unit (Neonatal Intensive Care Unit (NICU), the highest-level unit versus Special Care Baby Unit (SCBU) or Local Neonatal Unit (LNU), lower level units)). This was because it was anticipated that performance might differ by type of unit: data from the original study indicated that smaller units tended to have lower starting uptake levels, so more room for improvement compared to larger NICUs.

#### Sensitivity and subgroup analyses

As sensitivity analyses the above model was run on data aggregated to (a) the maternity-unit-level rather than the national level, and (b) the individual rather than national level. These two models additionally adjusted for type of maternity unit (NICU vs SCBU/LNU), regional clustering by AHSN, and were weighted on the number of eligible births per unit per month. Other sensitivity analyses included assessing (c) the impact of excluding a ‘fuzzy’ implementation start window of +/-2 months, to account for some units starting slightly earlier or later than their officially recorded start date; (d) the impact of excluding the final 2 months of data, due to concerns about completeness of the most recent data for some units; (e) the impact of using a longer pre-NPP comparison period of four years; and (f) the impact of including more mature pre-term babies up to 34 weeks’ gestation in the analysis. A sub-group analysis was performed on the 40 units that had participated in a connected study, an RCT nested within the main NPP(17), as their performance could plausibly differ from other maternity units.

### Economic evaluation

#### MgSO_4_ treatment cost-effectiveness

Economic analysis combines evidence of the treatment (MgSO_4_) effect with evidence of the implementation (NPP) effect(18). For the former, estimates of the cost-effectiveness of MgSO_4_ treatment were adopted from Bickford and colleagues’ results(6, 19, 20), with their estimates converted to GBP currency and 2019 prices (Supplementary Table 1). This analysis accounts for the health impacts and cost savings of MgSO_4_ administered in births <30 and <32 weeks’ gestation.

#### NPP implementation costs and effectiveness

The mean implementation cost per unit of the NPP was estimated from data supplied by the NPP team and PReCePT study team, and reported previously(12, 17). The total cost of the NPP at a national level was estimated at £936,747(12) (which includes the funded backfill of clinical time for NPP ‘champion’ midwives at each maternity unit, and regional support from AHSNs).

From the multivariable linear regression model described above, we estimated the NPP effectiveness as the difference between the predicted level of MgSO_4_ use over time compared to a counterfactual level of MgSO_4_ use, representing what may have occurred in the absence of the NPP, assuming a continuation of the pre-NPP trend in MgSO_4_ uptake. The main measure of NPP effectiveness was the area-between-the-curves. Primary analysis used a linear distribution to estimate the counterfactual based on the pre-NPP predicted trend. Sensitivity analysis used a beta distribution to estimate this counterfactual, to account for MgSO_4_ uptake as a proportion.

#### Policy cost-effectiveness analysis

The cost-effectiveness analysis was conducted from a societal lifetime perspective. Net monetary benefit (NMB) of the NPP was estimated over the four years since its launch, by combining analysis of the costs and effectiveness of the NPP with the lifetime societal cost, and health gains associated with MgSO_4_ treatment. This analysis used a framework previously developed to conduct economic evaluations of implementation initiatives(21). A willingness-to-pay threshold of £20,000 per Quality Adjusted Life Year (QALY) gained was used to determine NPP cost-effectiveness, following NICE guidelines(22). A positive NMB indicated that the implementation initiative was cost-effective. The net increment of the number of patients that received MgSO_4_, and the implementation cost-effectiveness per additional patient treated, was also estimated. The analysis used the area-between-the-curves estimate of NPP effectiveness with a linear counterfactual, and a sensitivity analysis using a beta counterfactual.

Probabilistic analysis was conducted using a Monte Carlo simulation with 10,000 samples drawn from parameter distributions. Point estimates, probabilistic distribution assumptions, and parameter source estimates are reported in Supplementary Table 2. Cost-effectiveness planes and cost-effectiveness acceptability curves were plotted for willingness-to-pay thresholds from zero to £100,000 per QALY gained for the policy cost-effectiveness of the NPP intervention.

#### Secondary economic analysis

As evidence on the lifetime cost-effectiveness of antenatal MgSO_4_ covers babies born up to 32 weeks’ gestation, cost-effectiveness analysis was performed only for babies <30 and <32 weeks’ gestation.

Statistical software Stata version 17 and R version 4.3.1 were used for all statistical analyses.

## RESULTS

### Baseline characteristics

In 2017, the year before NPP roll-out, a total of 4091 babies born under 30 weeks’ gestational age were admitted to neonatal units in England, 296 in Scotland, and 182 in Wales. The majority of births were in maternity units with a NICU (62.6% in England, 83.5% in Scotland, 63.0% in Wales). Other than the number of babies admitted, study populations were largely comparable across the three nations with respect to other covariates (Table 1).

**Table 1:**
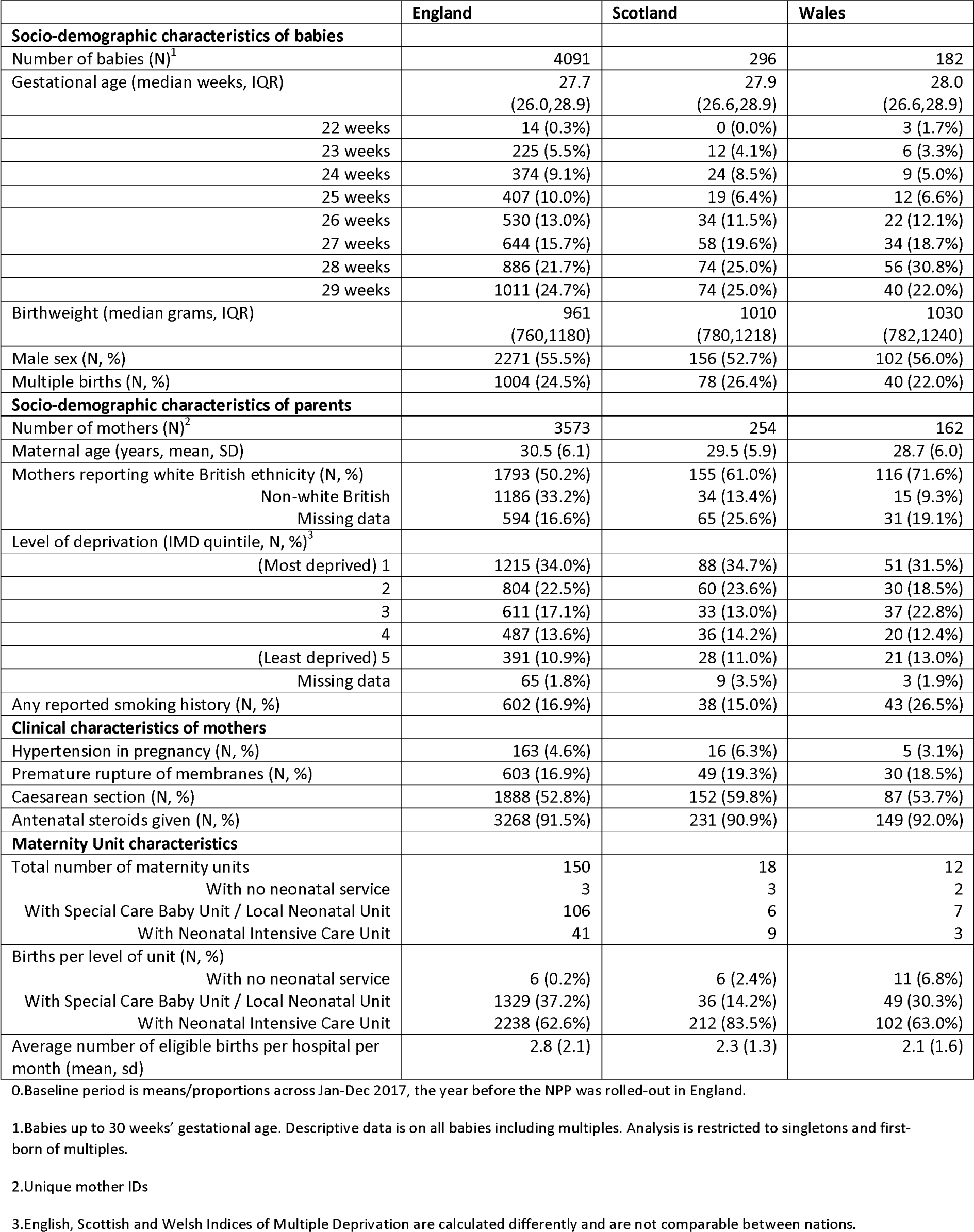
Baby, mother, and maternity unit characteristics by nation at baseline^0^.

|  | England | Scotland | Wales |
| --- | --- | --- | --- |
| **Socio-demographic characteristics of babies** |  |  |  |
| Number of babies (N)<sup>1</sup> | 4091 | 296 | 182 |
| Gestational age (median weeks, IQR) | 27.7<br>(26.0,28.9) | 27.9<br>(26.6,28.9) | 28.0<br>(26.6,28.9) |
| 22 weeks | 14 (0.3%) | 0 (0.0%) | 3 (1.7%) |
| 23 weeks | 225 (5.5%) | 12 (4.1%) | 6 (3.3%) |
| 24 weeks | 374 (9.1%) | 24 (8.5%) | 9 (5.0%) |
| 25 weeks | 407 (10.0%) | 19 (6.4%) | 12 (6.6%) |
| 26 weeks | 530 (13.0%) | 34 (11.5%) | 22 (12.1%) |
| 27 weeks | 644 (15.7%) | 58 (19.6%) | 34 (18.7%) |
| 28 weeks | 886 (21.7%) | 74 (25.0%) | 56 (30.8%) |
| 29 weeks | 1011 (24.7%) | 74 (25.0%) | 40 (22.0%) |
| Birthweight (median grams, IQR) | 961<br>(760,1180) | 1010<br>(780,1218) | 1030<br>(782,1240) |
| Male sex (N, %) | 2271 (55.5%) | 156 (52.7%) | 102 (56.0%) |
| Multiple births (N, %) | 1004 (24.5%) | 78 (26.4%) | 40 (22.0%) |
| **Socio-demographic characteristics of parents** |  |  |  |
| Number of mothers (N)<sup>2</sup> | 3573 | 254 | 162 |
| Maternal age (years, mean, SD) | 30.5 (6.1) | 29.5 (5.9) | 28.7 (6.0) |
| Mothers reporting white British ethnicity (N, %) | 1793 (50.2%) | 155 (61.0%) | 116 (71.6%) |
| Non-white British | 1186 (33.2%) | 34 (13.4%) | 15 (9.3%) |
| Missing data | 594 (16.6%) | 65 (25.6%) | 31 (19.1%) |
| Level of deprivation (IMD quintile, N, %)<sup>3</sup> |  |  |  |
| (Most deprived) 1 | 1215 (34.0%) | 88 (34.7%) | 51 (31.5%) |
| 2 | 804 (22.5%) | 60 (23.6%) | 30 (18.5%) |
| 3 | 611 (17.1%) | 33 (13.0%) | 37 (22.8%) |
| 4 | 487 (13.6%) | 36 (14.2%) | 20 (12.4%) |
| (Least deprived) 5 | 391 (10.9%) | 28 (11.0%) | 21 (13.0%) |
| Missing data | 65 (1.8%) | 9 (3.5%) | 3 (1.9%) |
| Any reported smoking history (N, %) | 602 (16.9%) | 38 (15.0%) | 43 (26.5%) |
| **Clinical characteristics of mothers** |  |  |  |
| Hypertension in pregnancy (N, %) | 163 (4.6%) | 16 (6.3%) | 5 (3.1%) |
| Premature rupture of membranes (N, %) | 603 (16.9%) | 49 (19.3%) | 30 (18.5%) |
| Caesarean section (N, %) | 1888 (52.8%) | 152 (59.8%) | 87 (53.7%) |
| Antenatal steroids given (N, %) | 3268 (91.5%) | 231 (90.9%) | 149 (92.0%) |
| **Maternity Unit characteristics** |  |  |  |
| Total number of maternity units | 150 | 18 | 12 |
| With no neonatal service | 3 | 3 | 2 |
| With Special Care Baby Unit / Local Neonatal Unit | 106 | 6 | 7 |
| With Neonatal Intensive Care Unit | 41 | 9 | 3 |
| Births per level of unit (N, %) |  |  |  |
| With no neonatal service | 6 (0.2%) | 6 (2.4%) | 11 (6.8%) |
| With Special Care Baby Unit / Local Neonatal Unit | 1329 (37.2%) | 36 (14.2%) | 49 (30.3%) |
| With Neonatal Intensive Care Unit | 2238 (62.6%) | 212 (83.5%) | 102 (63.0%) |
| Average number of eligible births per hospital per month (mean, sd) | 2.8 (2.1) | 2.3 (1.3) | 2.1 (1.6) |

0. Baseline period is means/proportions across Jan-Dec 2017, the year before the NPP was rolled-out in England.
1. Babies up to 30 weeks' gestational age. Descriptive data is on all babies including multiples. Analysis is restricted to singletons and first-born of multiples.
2. Unique mother IDs
3. English, Scottish and Welsh Indices of Multiple Deprivation are calculated differently and are not comparable between nations.

### Historical trends

In 2014, MgSO_4_ uptake was around 20% in England, 40% in Scotland, and 10% in Wales. Uptake improved over time in all three nations, with the rate of change slowing down at higher levels of treatment (ceiling effect). Although national levels appeared to converge in the latest 2022 data, there was visual suggestion that since the launch of the NPP, uptake may have been accelerated in England compared to the devolved nations. However, due to relatively smaller numbers, there was high variation in monthly uptake for Scotland and Wales, which limited formal assessment of parallel trends (Figure 1).

**Figure 1:**
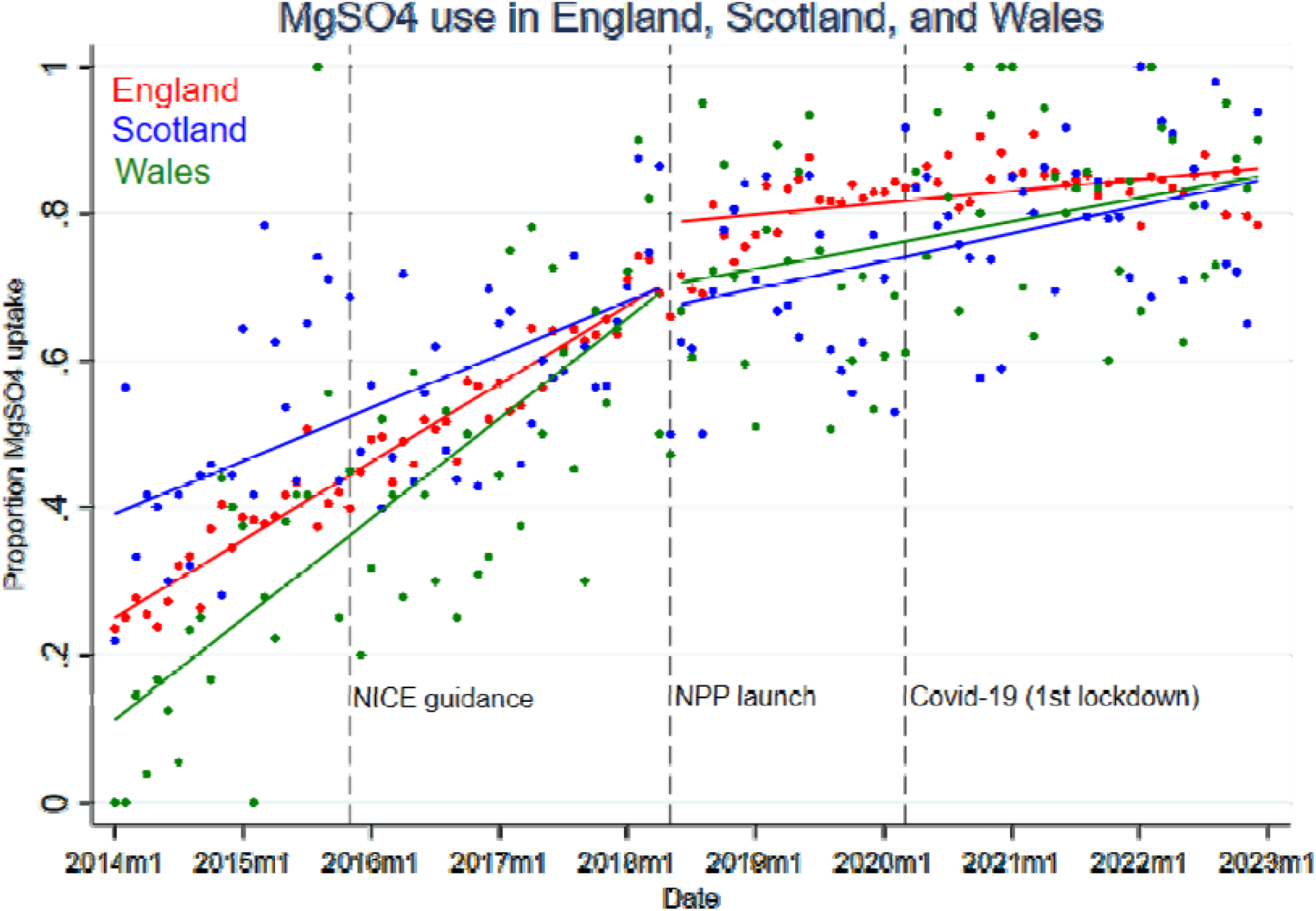
MgSO4 uptake in England, Scotland and Wales, 2014 to 2022.

### Pre/post-NPP comparison

In England, overall MgSO_4_ uptake rose from 65.8% in 2017 to 85.5% in 2022 (62.3% to 81.4% in Scotland, 61.6% to 86.6% in Wales). The amount of missing data fell from around 5% in 2017 to under 1% in 2022. Imminent delivery was the most commonly recorded reason for not giving MgSO_4_, accounting for around 15% of eligible babies in 2017, dropping to around 10% in England and Wales in 2022. The number recorded as not offered MgSO_4_ fell from around 7% to around 1% across all three nations (Supplementary Table 3).

### Estimate of NPP effectiveness

The adjusted model estimated an average 5.8 (95% CI 2.7 to 8.9, p<0.001) percentage point increase in MgSO_4_ uptake in England across the four years post-NPP, compared to the one year pre-NPP. Much of the gains appeared to take place as a step-change in the first two years of the programme. There were additional gains in years three and four (at which point the improvement became statistically significant), although confidence intervals overlap with estimates from the first two years (Table 2).

**Table 2:** Change in MgSO_4_ uptake in England from before to after the NPP^0^.

|  | Step-change in MgSO<sub>4</sub> uptake after intervention (percentage points)<sup>1</sup> | 95% CI | p-value | Change in slope post-intervention compared to pre-intervention | 95% CI | p-value | Overall slope post-intervention | 95% CI | p-value |
| --- | --- | --- | --- | --- | --- | --- | --- | --- | --- |
| Unadjusted<sup>2</sup> | 11.8 | 10.0 to 13.6 | <0.001 | - | - | - |  |  |  |
| Fully adjusted<sup>3</sup> | 5.8 | 2.7 to 8.9 | <0.001 | -0.87 | -1.18 to -0.57 | <0.001 | 0.02 | -0.06 to 0.10 | 0.622 |
| **Cumulative increase per-year across the follow-up period<sup>2</sup>** |  |  |  |  |  |  |  |  |  |
| Year 1 | 3.1 | -1.3 to 7.4 | 0.167 | -0.76 | -1.48 to -0.05 | 0.037 | 0.19 | -0.31 to 0.69 | 0.450 |
| Year 1-2 | 3.5 | -0.2 to 7.3 | 0.067 | -0.57 | -0.91 to -0.23 | <0.001 | 0.25 | 0.09 to 0.41 | 0.002 |
| Year 1-3 | 5.6 | 2.1 to 9.1 | 0.002 | -0.86 | -1.25 to -0.48 | <0.001 | 0.05 | -0.05 to 0.16 | 0.302 |
| Year 1-4 | 5.8 | 2.7 to 8.9 | <0.001 | -0.87 | -1.17 to -0.57 | <0.001 | 0.02 | -0.06 to 0.10 | 0.622 |
| **Secondary, sensitivity and sub-group analyses<sup>2</sup>** |  |  |  |  |  |  |  |  |  |
| Analysis at maternity-unit level<sup>4</sup> | 5.8 | 2.8 to 8.8 | <0.001 | -0.83 | -1.47 to -0.18 | 0.012 | 0.003 | -0.05 to 0.06 | 0.923 |
| Analysis at individual-level<sup>4</sup> | 5.9 | 2.5 to 9.3 | 0.001 | -0.76 | -1.17 to -0.34 | <0.001 | 0.02 | -0.03 to 0.07 | 0.449 |
| Excluding implementation start window (2 months each side of start) | 6.6 | 2.9 to 10.3 | <0.001 | -0.86 | -1.17 to -0.54 | <0.001 | -0.01 | -0.09 to 0.07 | 0.820 |
| Excluding final 2 months (?data quality) | 5.8 | 2.6 to 8.9 | <0.001 | -0.82 | -1.14 to -0.50 | <0.001 | 0.04 | -0.04 to 0.12 | 0.346 |
| Comparing 4 years pre with 4 years post | 6.0 | 3.7 to 8.4 | <0.001 | -0.74 | -0.82 to -0.65 | <0.001 | 0.01 | -0.07 to 0.08 | 0.895 |
| PReCePT cRCT units only (n=40) | 8.6 | 2.4 to 14.9 | 0.007 | -0.79 | -1.37 to -0.20 | <0.001 | 0.04 | -0.09 to 0.16 | 0.563 |
| Including babies up to 34 weeks gestation | 8.7 | 6.4 to 11.0 | <0.001 | -0.69 | -0.90 to -0.48 | <0.001 | 0.046 | -0.02 to 0.11 | 0.157 |

<sup>0</sup> All data on singletons and first born of multiples <30 weeks' gestation and admitted to an NHS Neonatal unit
<sup>1</sup> Percentage point difference in uptake between mean across the 12m pre-NPP, and mean across the four years post-NPP <sup>2</sup> Crude regression of uptake post-implementation compared with pre-implementation.
<sup>3</sup> Fully adjusted model: includes interaction between pre-post period and study month to capture change in slope as well as step-change. Adjusted for covariates as monthly aggregates: mean maternal age, proportion who identified as of white British ethnicity, mean IMD decile, proportion of multiple births, proportion with pregnancy hypertension, reported smokers, type of birth (c-section versus vaginal delivery), birthweight adjusted for gestational age as a z-score. Model weighted on unit size.
<sup>4</sup> Additionally adjusted for level of birth unit, and regional clustering by AHSN.

Estimates were robust to sensitivity analyses (Table 2). There was indication of greater improvement in MgSO_4_ use when including babies up to 34 weeks’ gestational age in the analysis (8.7 percentage point increase in MgSO_4_ uptake, 95%CI 6.4 to 11.0, p<0.001), and in the 40 units in the PReCePT RCT (8.6 percentage point increase in MgSO_4_ uptake, 95%CI 2.4 to 14.9, p=0.007). There was some evidence that lower-level maternity units improved more than higher-level units (SCBUs and LNUs: 9.1 percentage points change, 95%CI 3.9 to 14.3, p=0.001. NICUs: 4.1 percentage points change, 95%CI 0.6 to 7.6, p=0.022), reflecting the fact that NICUs tended to have higher starting levels.

### Impact of the COVID-19 pandemic

In 2020 there was a slight declining trend in MgSO_4_ use coinciding with the start of the COVID-19 pandemic, which continued to the end of the dataset at the end of 2022. The use of antenatal steroids (another, more well-established protective treatment for preterm babies) had an almost identical decline over this same period (Supplementary Figure 1).

### Economic evaluation

The impact of the NPP is illustrated in Figure 2, using a linear distribution (Figure 2a) and a beta distribution (Figure 2b) as counterfactuals. Probabilistic analysis estimated that the additional use of MgSO_4_ attributed to the NPP was equivalent to 3.0 percentage point improvement on average over seven months, which equates to an additional 64 of the 2136 pre-term (<30 weeks’ gestation) babies receiving treatment (Table 3). The lifetime and societal NMB of the NPP was about £597000, or £279 per preterm baby. The probability of the NPP being cost-effective was 89% (Table 3, Supplementary Figure 2).

**Figure 2:**
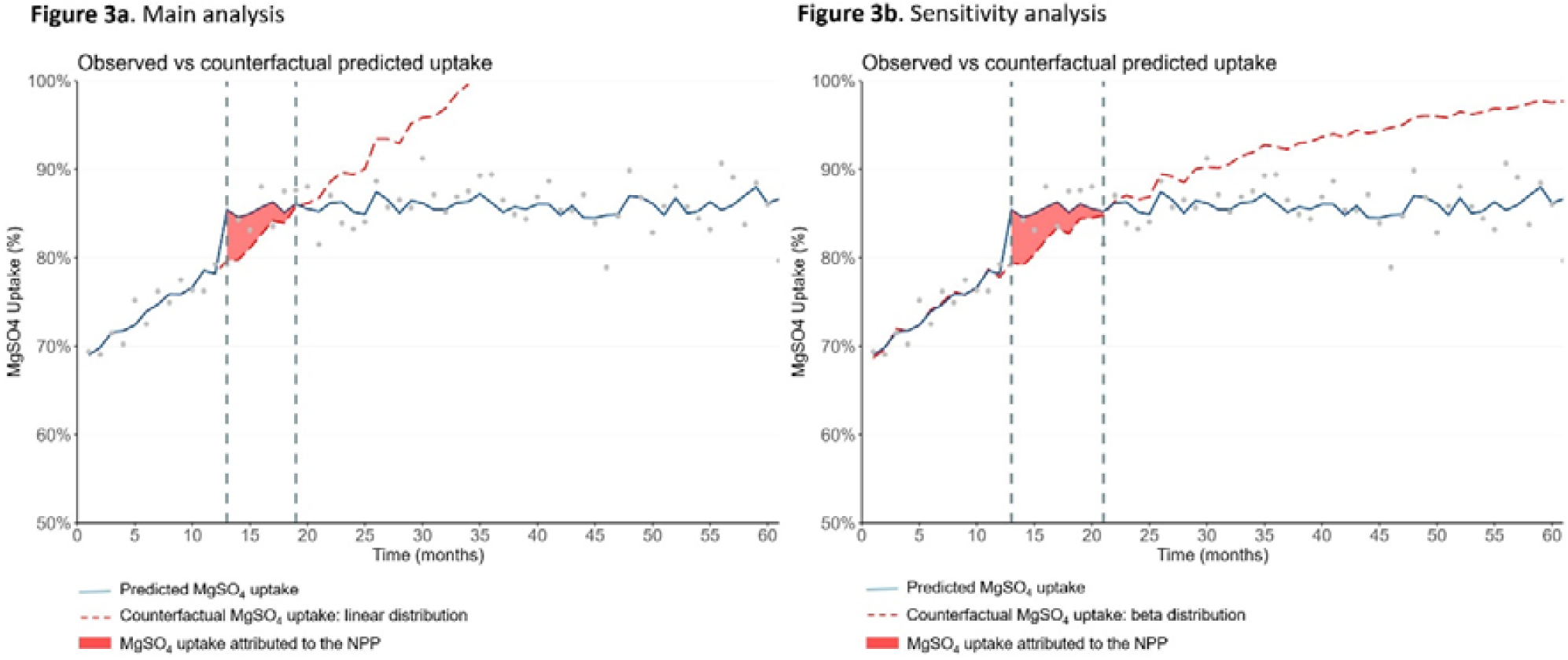
Predicted MgSO4 uptake, Counterfactual and Area-Between-Curves from Interrupted Time Series analyses.

**Table 3.** Probabilistic Cost-Effectiveness Results of the NPP from Interrupted Time Series Analysis (<30 weeks’ gestation)

|  | Main analysis<br>(linear distribution<br>counterfactual) | Sensitivity analysis<br>(beta distribution<br>counterfactual) |
| --- | --- | --- |
| Period of benefit, months | 7 | 10 |
| Number of pre-term babies ($\leq 3$ weeks), $N$ | 2136 | 3129 |
| Change in proportion of pre-term babies treated with $\text{MgSO}_4$, $\Delta b_i$, % | 3.0% (1.5%; 4.5%) | 2.9% (2.3%; 3.6%) |
| Net Increment of pre-term babies treated with $\text{MgSO}_4$, $\Delta pat$ | 64 (32; 97) | 92 (72; 112) |
| Net cost of implementation, $\Delta C_i$, £ | 936747 | 936747 |
| Implementation cos-effectiveness, $\Delta C_i / \Delta Pat$, £ per additional patient treated | 14576 (29284; 9669) | 10219 (13040; 8386) |
| Lifetime health effect of $\text{MgSO}_4$ treatment per patient, $\Delta bt$, QALY | 0.24 (0.16; 0.33) | 0.24 (0.16; 0.33) |
| Lifetime costs of $\text{MgSO}_4$ treatment per patient, $\Delta ct$, £ | -19064 (-13310; -25648) | -19064 (-13310; -25648) |
| Net Monetary Benefit of the Policy, $NMB_p$, £<sup>1</sup> | 596538 (-221748; 1541786) | 1251511 (558115; 2071244) |
| Probability of being cost-effective, % | 89% | 100% |

<sup>1</sup> At a willingness-to-pay threshold of £20,000 per QALY gained

The sensitivity analysis with the beta distribution counterfactual, estimated a longer period of impact (an additional three months) over which there was additional use of MgSO_4_ attributed to the NPP equivalent to a 2.9 percentage point improvement on average over 10 months, which equates to an additional 92 of 3129 pre-term babies receiving treatment (Figure 2b). Accounting for the total cost of the NPP, and the lifetime health gains and cost savings of MgSO_4_ treatment, the NMB was estimated to be £1.3m, or £400 per preterm baby. The probability of the NPP being cost-effective in this analysis was 100% (Table 3).

Expanding the analysis to include babies up to 32 weeks’ gestation estimated additional use of MgSO_4_ attributed to the NPP equivalent to a 4.4 percentage point improvement on average over nine months, which equates to an additional 215 pre-term babies treated. As the total cost of the NPP was fixed, and not sensitive to the number of babies treated, the NMB of the NPP was about £4.2m, or £853 per pre-term baby. The probability of NPP being cost-effective was 100% at the willingness-to-pay threshold of £20,000 per QALY gained (Supplementary Table 4 and Supplementary Figure 3). For the sensitivity analysis with the beta distribution the NMB of NPP was about £5.4m, or £700 per preterm baby. The probability of cost-effectiveness was 100% (Supplementary Table 4).

Expanding the analysis further again to include babies up to 34 weeks’ gestation shows that the additional use of MgSO_4_ attributed to the NPP was equivalent to 7.3 percentage point improvement on average over seven months. This means an additional 961 babies receiving treatment. Using a beta distribution for the counterfactual showed a 6.5 percentage point improvement attributed to the NPP over 12 months (698 additional patients). The NMB of the NPP was not calculated for this more mature group of preterm babies, as currently there is no available estimate for the cost-effectiveness of MgSO_4_ treatment for babies born above 31^+6^ weeks’ gestation.

## DISCUSSION

The original NPP evaluation found evidence of improved MgSO_4_ use over the first 12 months following implementation. This extended evaluation found that the improvements have largely been sustained over the first four years following implementation, although there was suggestion of a slight decline in use coinciding with the pandemic. The benefits applied both to the target population of births <30 weeks’ gestation, but also to more mature preterm babies up to 34 weeks’ gestation. The programme was associated with a net monetary benefit of about £0.6m for babies up to 29 weeks’ gestation, rising to about £4.2m when babies up to 31 weeks’ gestation are included. Compared to the devolved nations, uptake appeared to improve faster in England in the first two years following the NPP launch. By the end of 2022 however, the three nations were broadly comparable with delivery of MgSO_4_ to around 81-87% of eligible mothers. This is at the higher end of levels reported internationally (69% to 87%(23–26)) following guidelines or interventions to increase MgSO_4_ uptake.

Comparisons between England, Scotland, and Wales should be interpreted with caution: firstly because there is high variability due to small numbers in the devolved nations data, which limits formal statistical comparison of trends. Secondly because Scotland and Wales were implementing their own MgSO_4_ initiatives (e.g. MCQIC(27), PERIPrem Cymru(28)), complicating their position as a control group. Thirdly because there was also accessing of the English PReCePT toolkit and implementation resources during this time period (66 downloads from Wales, 32 from Scotland, 2018-2022, AHSN data, unpublished). This ‘contamination’ means that the boundaries of the target population are fuzzy, and again the devolved nations cannot be considered optimal controls. Finally, the three nations’ trends in uptake prior to the NPP were also not parallel, due to variation in starting levels in 2014, and this meant that formal statistical comparison of their improvements (for example including all three nations in an interrupted time series or difference in difference analysis) was not appropriate.

The COVID-19 pandemic could plausibly have impacted on MgSO_4_ use: the broad impact on staffing and quality of care affecting all parts of the NHS(29), and a specific impact on expecting mothers who may have presented at hospital later due to concerns about infection and giving birth alone, could both lead to missed opportunities to give MgSO_4_(30, 31). Analysis of future data will be important to explore what may be a temporary negative effect of the pandemic, versus what might be a natural waning of the initially positive effects of the NPP. From the observation that antenatal steroid use (historically well-established at high levels) declined almost identically to MgSO_4_ use over the pandemic period, the national decline may be associated with the pandemic. This finding is concerning and may illustrate the fragility of complex healthcare systems. Further follow up is important to monitor both metrics.

The observation of greater improvements in uptake when including more mature preterm births (all babies born up to 34 weeks’ gestation) is interesting. While the more mature preterm infants represent a much larger proportion of preterm births, interpretation is complex; their profile of underpinning antenatal disease, and hence their presentation to healthcare, may vary from the more extreme preterm presentation. This could be expected to impact on the subsequent ease of delivering antenatal MgSO_4_. Evidence on the protective effect of MgSO_4_ in babies 30-34 weeks’ is less clear(3, 32, 33) and although long term neurological impacts remain higher in these groups than term-born peers(34), due to their greater numbers, even small shifts in risks may have substantial population benefits(35).

It is likely to be more difficult to improve from 85% to 90% uptake, compared to improving from 65% to 70% uptake. Further overall increases in MgSO_4_ use may be a challenge without concerted effort at the lower-performing units. However, as some units do report higher (>90%) uptake (perhaps through better triaging and monitoring of symptoms), we argue that their performance should be used as the benchmark for quality of care, and investment in supporting the lower-performing units is likely to be cost-effective.

The creation of clinical guidelines alone is often not enough to ensure that evidence-based interventions become standard practice. A relevant example here is the case of antenatal steroids, which in the absence of a programme dedicated to getting this evidence into practice, took several decades for their use to become standard care. In contrast, and in the context of the NPP, the same improvements in use of MgSO_4_ were achieved within a few years.

### Strengths and limitations

This evaluation included effectiveness and cost-effectiveness analysis to capture the impact of the NPP implementation. These types of analysis have been highlighted as key components of implementation science research(18, 36). Results from the main and sensitivity analyses were consistent. Data covered a period of eight years, giving adequate time for analysis of trends. The study benefitted from high quality, routinely collected, national, longitudinal patient-level data. The key advantages of this comprehensive real-world data is that it provides high generalisability (included all maternity units in England, Scotland, and Wales, reflecting the nationwide situation), shows effectiveness in real-world conditions, and is less vulnerable to some biases such as recall, observer, and attrition bias. A limitation is that this sort of data does not necessarily include all the covariates of interest, and data quality and completeness is not always consistent across all sites.

A key limitation is that residual confounding cannot be excluded. We have tried to minimise the impact of confounding through robust analytic methods, and interpret findings with caution. In addition to the PReCePT programme, there are other factors that likely will have impacted on uptake, including the publication of definitive evidence on the protective effective of MgSO_4_ in 2009(37) MgSO_4_ use being reliably recorded as a Neonatal Data Analysis Unit (NDAU) audit quality metric for maternity units in 2014-15, and its use becoming a formal recommendation in the NICE Guidance in 2015(2). Other factors related to PReCePT include the original pilot study publishing positive results in 2017; and discussions with unit leads about the proposed NPP in 2017. The National Neonatal Audit Programme (NNAP) concluded in their report on 2020 data that “This rapid improvement [in MgSO_4_ use], particularly seen in England, is likely to result from the targeted approach of the PReCePT quality improvement initiative.”(38)

Another limitation is that the population investigated here included liveborn babies admitted to a neonatal unit, rather than the total population of mothers eligible for MgSO4. The rate of MgSO_4_ uptake may be different between these two populations, although any health benefit would only be realised in those investigated in this work. It would be an advantage if future research could explore, and compare, uptake in both populations, and the survival and neurodevelopmental outcomes for those preterm babies who did, and did not, receive MgSO_4_.

The Health Foundation and Health Data Research UK recently listed PReCePT as a case study model for a Learning Health System (i.e. a systematic approach to iterative, data-driven quality improvement(39)) and we propose that the PReCePT model could be used as an implementation blueprint for other quality improvement initiatives.

## CONCLUSIONS

Implementation of the National PReCePT Programme has plausibly helped accelerate uptake of MgSO_4_ in England, improving maternal and neonatal care, and positively impacting society in terms of direct patient benefit and future cost savings. Failure to deliver MgSO_4_ to eligible mothers should be considered inadequate care, and not financially sustainable for the NHS. MgSO_4_ as a quality metric should continue to be closely monitored, and further intervention may be warranted to achieve optimal treatment levels. Future research should quantify the patient outcomes in this same population, specifically the cases of cerebral palsy prevented, associated with the improvements in use of MgSO_4_.

## Data Availability

Anonymised individual-level data for this study are from the NNRD. Our data sharing agreement with the NNRD prohibits sharing data extracts outside of the University of Bristol research team. The NNRD data dictionary is available online and copies of the Statistical analysis plan are available at the Open Science Framework repository (https://osf.io/be76s/).

## Abbreviations

CP: Cerebral Palsy
MgSO_4_: Magnesium Sulphate
NDAU: Neonatal Data Analysis Unit
NMB: Net Monetary Benefit
NNRD: National Neonatal Research Database
NPP: National PReCePT Programme
PReCePT: Prevention of Cerebral Palsy in PreTerm Labour
QI: Quality Improvement

## Notes

### Competing Interest Statement

The authors have declared no competing interest.

### Funding Statement

This study was jointly funded by The Health Foundation (funder's reference 557668), the National Institute for Health and Care Research Applied Research Collaboration West (NIHR ARC West, core NIHR infrastructure funded: NIHR200181), and Health Innovation West of England (formerly the West of England Academic Health Science Network). The views expressed are those of the authors and not necessarily those of NHS England, NHS Improvement, the NIHR or the Department of Health and Social Care.

### Author Declarations

The PReCePT Programme Evaluation was granted a favourable ethical opinion by the UK National Health Service Health Research Authority (HRA project ID: 260504) and the University of Bristol Faculty of Health Sciences Research Ethics Committee (FREC Ref: 84582).

