## Supplementary materials for "Implementation of national guidelines on antenatal magnesium sulfate for neonatal neuroprotection in England, Scotland and Wales: Extended evaluation of the effectiveness and cost-effectiveness of the National PReCePT Programme"

### Supplementary file

#### Supplementary table 1: Estimated lifetime costs and QALYs per patient associated with MgSO4 treatment (2019 prices)

| Type of birth | Perspective | $MgSO_4$ | Cost, £ | Δcost, £ | QALYs | ΔQALYs |
|---|---|---|---|---|---|---|
| Imminent | Societal | Yes | 61971 | -23690 | 26.6 | 0.3 |
| | | No | 85661 | | 26.3 | |
| Threatened | Societal | Yes | 44068 | -15964 | 26.7 | 0.2 |
| | | No | 60032 | | 26.5 | |
| **Combined (40% imminent)** | **Societal** | **Yes** | **51229** | **-19054** | **26.7** | **0.24** |
| | | **No** | **70284** | | **26.4** | |

Based on Bickford et al. 2013

**Supplementary table 2: Point estimates, probability distributions, and source of parameter estimates used in the probabilistic analysis**

| Analysis | Element | Statistics | Linear | Distribution | Source |
|---|---|---|---|---|---|
| **Common estimates** | Health utility, QALYs | Mean (SE *) | 0.24 (0.05) | Beta distribution | Bickford et al. |
| | Lifetime costs, £ | Mean (SE *) | -19054 (-3811) | Gamma distribution | Bickford et al. |
| | Cost NPP, £ | Total | 936747 | N/A | PReCePT Study – Cost analysis |
| **Linear ITS >30 weeks gestation** | Pre-term babies, *N* | Sum | 2136 | N/A | NNRD data |
| | Change in the proportion of MgSO4 | Mean (SE) | 3.0% (0.9%) | Normal distribution | Area-Between-Curves |
| **Beta ITS >30 weeks gestation** | Pre-term babies, *N* | Sum | 3129 | N/A | NNRD data |
| | Change in the proportion of MgSO4 | Mean (SE) | 3.0% (0.4%) | Normal distribution | Area-Between-Curves |
| **Linear ITS >32 weeks gestation** | Pre-term babies, *N* | Sum | 4923 | N/A | NNRD data |
| | Change in the proportion of MgSO4 | Mean (SE) | 3.4% (1.0%) | Normal distribution | Area-Between-Curves |
| **Beta ITS >32 weeks gestation** | Pre-term babies, *N* | Sum | 7768 | N/A | NNRD data |
| | Change in the proportion of MgSO4 | Mean (SE) | 3.4% (0.3%) | Normal distribution | Area-Between-Curves |

*Standard errors are calculated as the 20% of the point estimates as in Bickford et al. 2013

#### Supplementary table 3: MgSO4 uptake in England, Scotland, and Wales, pre- and post-NPP[0]

| | England | | Scotland | | Wales | |
|---|---|---|---|---|---|---|
| | 2017[1] | 2022[2] | 2017[1] | 2022[2] | 2017[1] | 2022[2] |
| Total number of eligible births[0] | 3573 | 3286 | 254 | 253 | 162 | 135 |
| Total number of mothers **given** $MgSO_4$ (%) | 2223 (62.2%) | 2786 (84.8%) | 149 (58.7%) | 205 (81.0%) | 93 (57.4%) | 116 (85.9%) |
| Total number of mothers **not given** $MgSO_4$ (%) | 1158 (32.4%) | 474 (14.4%) | 90 (35.6%) | 47 (18.6%) | 58 (35.8%) | 18 (13.3%) |
| Total number with **missing** $MgSO_4$ data (%) | 192 (5.4%) | 26 (0.8%) | 15 (5.9%) | 1 (0.4%) | 11 (6.8%) | 1 (0.7%) |
| **$MgSO_4$ uptake[3]** (sd) | 65.8% (0.5) | 85.5% (0.4) | 62.3% (0.5) | 81.4% (0.4) | 61.6% (0.5) | 86.6% (0.3) |
| Reason $MgSO_4$ not given (%)[4] | | | | | | |
| Contraindicated | 11 (0.3%) | 5 (0.2%) | 2 (0.8%) | 1 (0.4%) | 0 (0.0%) | 1 (0.7%) |
| Declined | 3 (0.1%) | 3 (0.1%) | 0 (0.0%) | 1 (0.4%) | 1 (0.6%) | 0 (0.0%) |
| Delivery imminent | 548 (15.3%) | 329 (10.0%) | 42 (16.5%) | 38 (15.0%) | 23 (14.2%) | 13 (9.6%) |
| Not appropriate | 126 (3.5%) | 23 (0.7%) | 14 (5.5%) | 1 (0.4%) | 12 (7.4%) | 0 (0.0%) |
| Not offered | 240 (6.7%) | 32 (1.0%) | 18 (7.1%) | 2 (0.8%) | 14 (8.6%) | 2 (1.5%) |
| Data missing | 230 (6.4%) | 82 (2.5%) | 14 (5.5%) | 4 (1.6%) | 8 (4.9%) | 2 (1.5%) |

0. All data on singletons and first born of multiples <30 weeks' gestation and admitted to an NHS Neonatal unit

1. Across the year Jan-Dec 2017

2. Across the year Jan-Dec 2022

3. Uptake percentage calculated excluding missing values from the denominator, to fit with national audit reporting practices.

4. Percentage calculated out of total cases

**Supplementary table 4: Probabilistic Cost-Effectiveness Results of the NPP from Interrupted Time Series Analysis (<32 weeks' gestation)**

| | **Main analysis** (linear distribution counterfactual) | **Sensitivity analysis** (beta distribution counterfactual) |
|---|---|---|
| Period of benefit, months | 9 | 12 |
| Number of pre-term babies (≤ 3 weeks), *N* | 4923 | 7768 |
| Change in proportion of pre-term babies treated with $MgSO_4$, $\Delta b_i$, % | 4.4% (2.7%; 6.0%) | 3.4% (3.0%; 3.9%) |
| Net Increment of pre-term babies treated with MgSO4, $\Delta pat$ | 215 (133; 297) | 267 (230; 303) |
| Net cost of implementation, $\Delta C_i$, £ | 936747 | 936747 |
| Implementation cos-effectiveness, $\Delta Ci$ / $\Delta Pat$, £ per additional patient treated | 4350 (7031; 3153) | 3508 (4065; 3087) |
| Lifetime health effect of MgSO4 treatment per patient, $\Delta bt$, QALY | 0.24 (0.16; 0.33) | 0.24 (0.16; 0.33) |
| Lifetime costs of MgSO4 treatment per patient, $\Delta ct$, £ | -19064 (-13310; -25648) | -19064 (-13310; -25648) |
| Net Monetary Benefit of the Policy, $NMB_P$, £[1] | 4199799 (1986016; 6803391) | 5433488 (3668040; 7508699) |
| Probability of being cost-effective, % | 100% | 100% |

[1] At a willingness-to-pay threshold of £20,000 per QALY gained

**Supplementary Figure 1:**

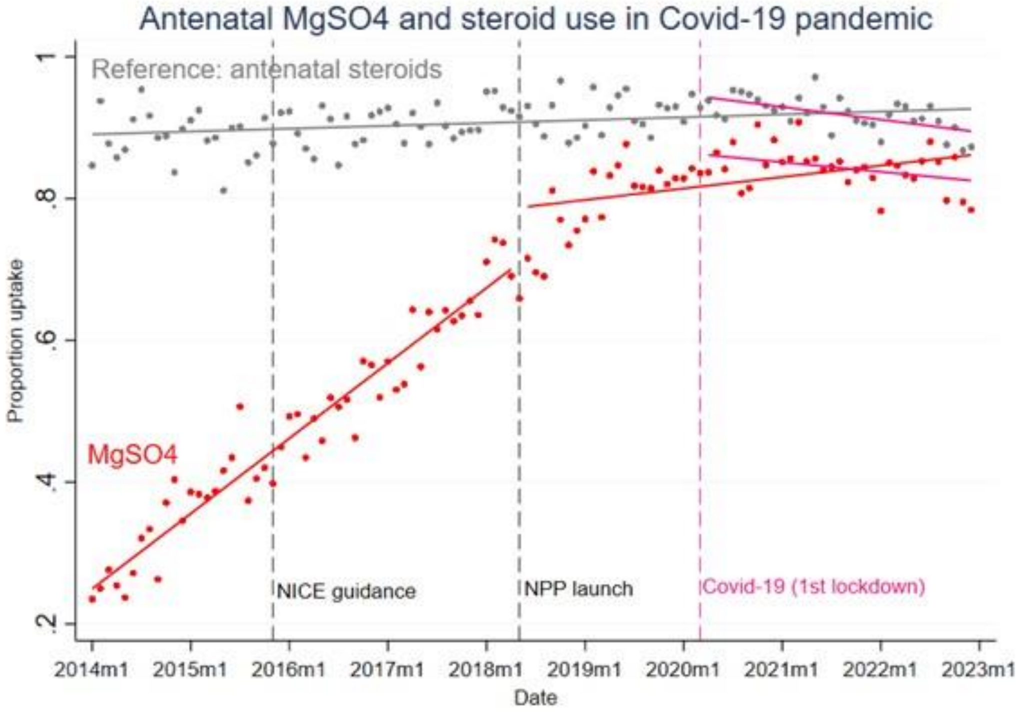

#### Supplementary Figure 2: Cost-Effectiveness Plane and Cost-Effectiveness Acceptability Curve from Linear Interrupted Time Series analysis (<30 weeks gestation)

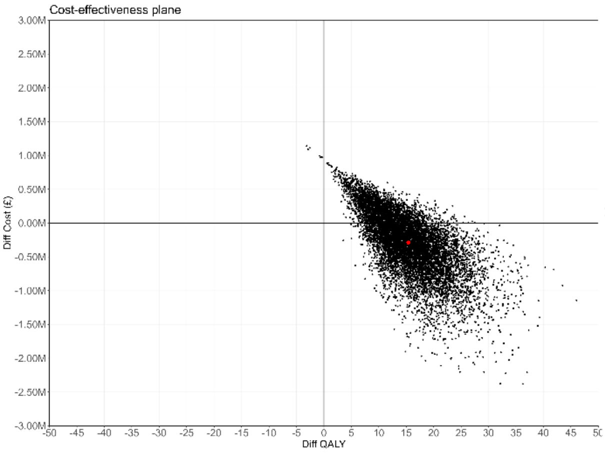


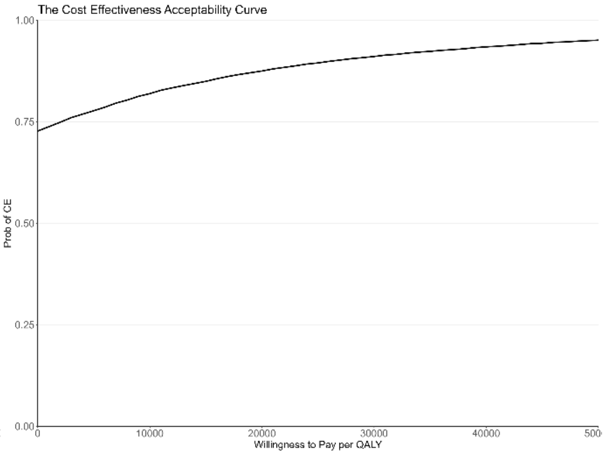


The graph on the left displays the results of Monte Carlo simulations with 10,000 iterations using the value ranges and distributions presented in Appendix 2. The horizontal axis represents the effect measures in Quality Adjusted Life Years (QALYs) for the National PReCePT Programme, and the vertical axis represents the cost. Datapoints falling in the top right quadrant indicate that the National PReCePT Programme was effective and costly. Datapoints falling bottom right quadrant indicate that the National PReCePT Programme was effective and cost-saving.

**Supplementary Figure 3: Cost-Effectiveness Plane and Cost-Effectiveness Acceptability Curve from Linear Interrupted Time Series analysis (<32 weeks gestation)**

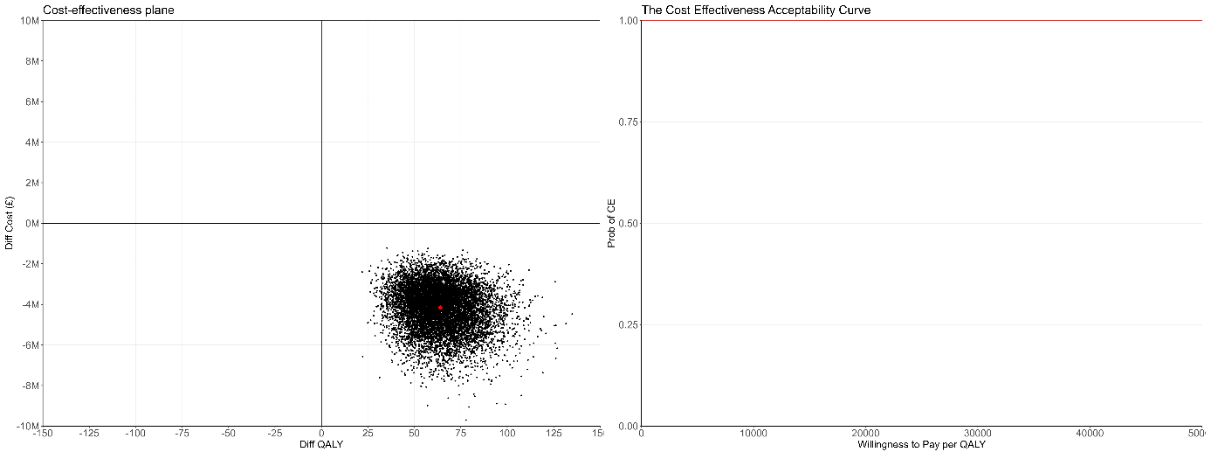


The graph on the left displays the results of Monte Carlo simulations with 10,000 iterations using the value ranges and distributions presented in Appendix 2. The horizontal axis represents the effect measures in Quality Adjusted Life Years (QALYs) for the National PReCePT Programme, and the vertical axis represents the cost. Datapoints falling bottom right quadrant indicate that the National PReCePT Programme was effective and cost-saving.
